# Artificial Intelligence in Reminiscence Therapy for Older Adults: A Systematic Review Protocol

**DOI:** 10.1101/2025.09.21.25336299

**Authors:** Ravi Shankar, Jaminah Ali, Xu Qian

## Abstract

**Background:** The global aging population faces increasing challenges related to cognitive decline, social isolation, and psychological well-being. Reminiscence therapy has emerged as an effective non-pharmacological intervention, and recent advances in artificial intelligence offer opportunities to enhance these therapeutic approaches.

**Objective:** To systematically review and synthesize evidence on the integration of artificial intelligence technologies in reminiscence therapy for aging populations, examining effectiveness, acceptability, implementation factors, and ethical considerations.

**Methods:** This protocol follows PRISMA-P guidelines and employs the PICOS framework. Comprehensive searches will be conducted in PubMed, Web of Science, Embase, CINAHL, MEDLINE, The Cochrane Library, PsycINFO, and Scopus from inception to October 2025. Studies examining AI-enhanced reminiscence therapy in adults aged 60 years and above will be included. Two independent reviewers will screen studies using Covidence, extract data, and assess quality using appropriate risk of bias tools. Data synthesis will involve meta-analysis where appropriate or narrative synthesis following SWiM guidelines.

**Discussion:** This protocol establishes a rigorous methodology for examining an emerging field at the intersection of gerontology and artificial intelligence. The systematic approach will provide evidence-based insights for clinical practice, technology development, and policy-making while identifying research gaps and ethical considerations in AI-enhanced therapeutic interventions for older adults.

## Introduction

The demographic transition toward an aging global population represents one of the most significant challenges facing healthcare systems worldwide. By 2050, the World Health Organization projects that approximately two billion people will be aged sixty years or older, with the most rapid growth occurring in low- and middle-income countries (1). This demographic shift necessitates innovative approaches to address the complex health and social needs of older adults, particularly regarding cognitive health, psychological well-being, and social connectedness. Among various non-pharmacological interventions developed for aging populations, reminiscence therapy has demonstrated consistent efficacy in improving mood, reducing depressive symptoms, and enhancing quality of life (2).

Reminiscence therapy involves the systematic recollection and sharing of personal memories and life experiences, typically facilitated through structured or semi-structured activities using prompts such as photographs, music, or familiar objects (3, 4). Traditional reminiscence therapy has shown moderate effect sizes for depression reduction and small to moderate effects for life satisfaction and emotional well-being in meta-analyses encompassing over one hundred controlled studies (5, 6). However, conventional delivery methods face limitations including resource intensiveness, accessibility barriers, inconsistent availability of trained facilitators, and challenges in maintaining therapeutic engagement over extended periods (7).

The emergence of artificial intelligence technologies presents unprecedented opportunities to address these limitations while potentially enhancing therapeutic outcomes. Artificial intelligence, encompassing machine learning, natural language processing, computer vision, and related computational approaches, offers capabilities for personalizing therapeutic experiences, automating content curation, and providing continuous support beyond traditional clinical settings (8-10). Recent developments include natural language processing systems that facilitate conversational interactions about past experiences, computer vision technologies that automatically organize personal photographs for memory prompting, and virtual reality systems that create immersive reminiscence experiences (11, 12).

Despite growing interest in AI-enhanced reminiscence therapy, the current evidence base remains fragmented across multiple disciplines including gerontology, psychology, computer science, and human-computer interaction. No comprehensive systematic review has synthesized findings regarding effectiveness, acceptability, implementation factors, and ethical considerations of AI-enhanced reminiscence interventions for older adults. This protocol addresses this gap by establishing a rigorous methodology for systematically examining the intersection of artificial intelligence and reminiscence therapy in aging populations, aiming to inform evidence-based practice, guide technology development, and establish priorities for future research.

## Methods

### Study Design and Framework

This systematic review protocol follows the Preferred Reporting Items for Systematic Review and Meta-Analysis Protocols (PRISMA-P) guidelines and has been registered with PROSPERO. The review employs the PICOS framework (Population, Intervention, Comparison, Outcomes, Study design) as the most appropriate structure for this clinical intervention review, providing comprehensive criteria for study selection while maintaining clarity and reproducibility (13).

### Eligibility Criteria

The PICOS framework guides our inclusion and exclusion criteria as detailed in Table 1. The population includes adults aged sixty years and above across all cognitive statuses, including cognitively healthy individuals, those with mild cognitive impairment, and individuals with dementia. No restrictions will be placed on setting, allowing inclusion of community-dwelling, assisted living, and institutionalized older adults. The intervention must involve reminiscence therapy incorporating artificial intelligence technologies, including but not limited to machine learning algorithms, natural language processing, computer vision, virtual agents, chatbots, or AI-powered virtual/augmented reality systems. Studies examining basic digital technologies without AI capabilities will be excluded. Comparison conditions may include traditional reminiscence therapy, usual care, waiting list controls, or alternative interventions. Studies without comparators will be included if they provide relevant implementation or feasibility data. Outcomes encompass cognitive function, psychological well-being, social engagement, quality of life, and technology-related measures such as usability and acceptability. All empirical study designs will be considered, including randomized controlled trials, quasi-experimental studies, observational studies, qualitative investigations, and mixed-methods research. Case reports, opinion pieces, and conference abstracts without full texts will be excluded.

**Table 1:** Inclusion and Exclusion Criteria.

| Criterion | Inclusion | Exclusion |
| --- | --- | --- |
| Population | Adults aged ≥60 years; All cognitive statuses; Any residential setting | Age <60 years; Non-human studies |
| Intervention | Reminiscence therapy with AI components (ML, NLP, computer vision, virtual agents, AI-powered VR/AR) | Basic digital tools without AI; AI for assessment only without therapeutic component |
| Comparison | Any comparator or no comparator | None |
| Outcomes | Cognitive, psychological, social, QoL, technology-related outcomes | None |
| Study Design | RCTs, quasi-experimental, observational, qualitative, mixed-methods, feasibility studies | Case reports, editorials, opinion pieces, abstracts only |
| Publication | Peer-reviewed journals/book chapters; Any language; Inception to October 2025 | Non-peer-reviewed sources (except dissertations) |

### Search Strategy

Comprehensive searches will be conducted across eight electronic databases: PubMed, Web of Science, Embase, CINAHL, MEDLINE, The Cochrane Library, PsycINFO, and Scopus, from each database’s inception to October 2025. The search strategy was developed in consultation with an information specialist and combines controlled vocabulary terms with free-text keywords across three concept areas: aging population, reminiscence therapy, and artificial intelligence. The search string has been adapted from established systematic review methodologies and tailored for this review’s specific focus:

((‘reminiscence therap^*^‘ OR ‘life review’ OR ‘life story’ OR ‘autobiographical memory’ OR ‘memory therap^*^‘ OR ‘narrative therap^*^‘ OR ‘personal histor^*^‘ OR ‘memory stimulation’ OR ‘biographical work’) AND (‘artificial intelligence’ OR ‘machine learning’ OR ‘deep learning’ OR ‘neural network^*^‘ OR ‘natural language processing’ OR ‘NLP’ OR ‘computer vision’ OR ‘chatbot^*^‘ OR ‘virtual agent^*^‘ OR ‘intelligent system^*^‘ OR ‘algorithm^*^‘ OR ‘automat^*^‘ OR ‘virtual reality’ OR ‘augmented reality’ OR ‘VR’ OR ‘AR’ OR ‘conversational AI’ OR ‘recommendation system^*^‘ OR ‘personali^*^ technolog^*^‘) AND (‘older adult^*^‘ OR ‘elderly’ OR ‘aging’ OR ‘ageing’ OR ‘aged’ OR ‘senior^*^‘ OR ‘geriatric^*^‘ OR ‘older people’ OR ‘elder^*^‘ OR ‘late life’))

Supplementary search strategies include hand-searching reference lists of included studies and relevant systematic reviews, forward citation tracking using Google Scholar, searching grey literature through ProQuest Dissertations and OpenGrey, and contacting experts in gerontechnology and digital health. No language restrictions will be applied, with translation services utilized when necessary.

### Study Selection

Study selection will be managed using Covidence systematic review software, which facilitates independent screening, tracks reviewer decisions, and automatically identifies conflicts requiring resolution. Following de-duplication, two independent reviewers will conduct title and abstract screening against predetermined eligibility criteria. Studies meeting inclusion criteria or those with insufficient information will proceed to full-text review. Full-text articles will be independently assessed by two reviewers, with specific reasons for exclusion documented. Disagreements at both stages will be resolved through discussion, with a third reviewer consulted if consensus cannot be reached. Inter-rater reliability will be calculated using Cohen’s kappa coefficient, with values above 0.80 considered excellent agreement. The PRISMA flow diagram will document the selection process, including reasons for exclusion at the full-text stage.

### Data Extraction

A standardized data extraction form will be developed and piloted on a sample of included studies before full implementation. Two independent reviewers will extract data using Covidence’s extraction templates, with discrepancies resolved through discussion and verification against original sources. Extracted data will encompass study characteristics including authors, publication year, country, setting, design, sample size, and funding sources; population characteristics including demographics, cognitive status, living arrangements, and technology experience; intervention details including AI technology specifications, reminiscence therapy protocols, duration, frequency, delivery mode, and training requirements; outcome measures including instruments used, timing of assessments, and statistical analyses; and implementation factors including feasibility indicators, adherence metrics, technical challenges, and resource requirements.

### Risk of Bias Assessment

Risk of bias will be assessed using design-appropriate tools by two independent reviewers. Randomized controlled trials will be evaluated using the Cochrane Risk of Bias tool version 2 (RoB 2), which assesses five domains: bias arising from the randomization process, deviations from intended interventions, missing outcome data, measurement of the outcome, and selection of the reported result (14). Each domain receives a judgment of low risk, some concerns, or high risk, contributing to an overall risk assessment. Non-randomized quantitative studies will be assessed using the Risk of Bias in Non-randomized Studies of Interventions (ROBINS-I) tool, evaluating seven domains including confounding, selection bias, intervention classification, deviations from intended interventions, missing data, outcome measurement, and selective reporting (15).

Qualitative studies will be evaluated using the Critical Appraisal Skills Programme (CASP) Qualitative Checklist (16), examining ten criteria related to research aims, methodology, design appropriateness, recruitment, data collection, researcher reflexivity, ethical considerations, analytical rigor, findings clarity, and research value. Mixed-methods studies will be assessed using the Mixed Methods Appraisal Tool (MMAT), which evaluates qualitative components, quantitative components, and method integration. Risk of bias assessments will inform sensitivity analyses and GRADE evaluations of evidence quality (17). Disagreements in risk of bias judgments will be resolved through discussion, with arbitration by a third reviewer when necessary.

### Data Synthesis

The data synthesis strategy will employ a convergent integrated approach that enables parallel synthesis of quantitative and qualitative evidence with subsequent integration to comprehensively address review objectives. For quantitative synthesis, clinical and methodological heterogeneity will be systematically assessed to determine the appropriateness of statistical pooling. Where sufficient homogeneity exists (I^2^ <75% with clinical similarity), meta-analysis will be conducted using random-effects models (DerSimonian-Laird or restricted maximum likelihood estimation) to account for between-study variance (18). Effect sizes will be calculated as standardized mean differences (Hedges’ g) for continuous outcomes and risk ratios for dichotomous outcomes, with 95% confidence intervals and prediction intervals to indicate the range of true effects in future studies. Forest plots will visualize individual study contributions and pooled estimates. When meta-analysis is inappropriate due to substantial heterogeneity, narrative synthesis will follow the Synthesis Without Meta-analysis (SWiM) framework, with studies grouped by meaningful characteristics such as intervention type (conversational AI versus VR-based reminiscence), population cognitive status, or setting. Within groups, patterns of effects will be described using consistent terminology, direction of effect plots, and vote counting based on direction of effect rather than statistical significance.

Qualitative synthesis will employ thematic synthesis methods adapted from Thomas and Harden (2008) (19), involving three analytical stages to capture implementation experiences, user perspectives, and contextual factors. Line-by-line coding of findings sections from included qualitative studies will identify concepts relevant to AI-enhanced reminiscence therapy implementation, with particular attention to user experiences with different AI technologies, acceptability factors across population groups, implementation challenges and solutions, and ethical considerations. Descriptive themes will be developed through hierarchical grouping of codes, maintaining close alignment with primary study findings, while analytical themes will be generated through interpretation that goes beyond individual studies to address review questions about optimal implementation strategies and contextual factors influencing success. The synthesis will follow ENTREQ (Enhancing Transparency in Reporting the synthesis of Qualitative research) guidelines, maintaining a clear audit trail from data extraction through coding to theme development, with exemplar quotes illustrating key themes. Mixed-methods studies will have their quantitative and qualitative components extracted and synthesized with respective study types, with additional attention to how integration was achieved and what insights emerged from the mixed-methods approach.

Integration of quantitative and qualitative syntheses will follow a convergent design where separate syntheses are conducted in parallel before integration through joint displays, matrices, and narrative weaving to identify convergence, complementarity, and divergence between evidence types. A matrix will map qualitative themes against quantitative outcomes to explore relationships; for instance, qualitative findings about technology anxiety may explain heterogeneity in quantitative engagement metrics, while implementation themes will be examined against effectiveness outcomes to identify potential implementation-effectiveness relationships. Configuration analysis will explore whether certain combinations of intervention features (type of AI, level of personalization), population characteristics (cognitive status, technology experience), and contextual factors (setting, support available) are associated with better outcomes. This integrated synthesis will address questions that neither quantitative nor qualitative evidence alone can answer, such as why certain AI-enhanced interventions work better for specific populations or how implementation context influences effectiveness.

Subgroup analyses and sensitivity analyses will explore sources of heterogeneity and assess the robustness of findings across key variables determined a priori. Planned subgroups include cognitive status categories, types of AI technology, delivery settings, intervention intensity, level of human support, and participant characteristics including age groups, gender, and technology experience, with analyses conducted only when sufficient studies are available (minimum 10 studies with at least 3 per subgroup). Sensitivity analyses will systematically vary inclusion decisions and analytical approaches, including excluding studies at high risk of bias, removing grey literature, examining outlier influence, and testing different meta-analytical models. The certainty of evidence will be assessed using GRADE for quantitative outcomes and CERQual for qualitative findings, providing transparent evaluation of confidence in synthesized evidence. Publication bias will be assessed through funnel plots and Egger’s test where appropriate (≥10 studies), comparison of published versus grey literature effects, and searching trial registries for unpublished studies. The synthesis will produce actionable findings that inform clinical practice, technology development, and policy decisions while clearly communicating the strength and limitations of available evidence.

## Discussion

This systematic review protocol addresses a critical gap in the literature by establishing a rigorous methodology for synthesizing evidence on AI-enhanced reminiscence therapy for aging populations. The protocol’s comprehensive approach, utilizing the PICOS framework and extensive database searching from inception to October 2025, ensures thorough coverage of this rapidly evolving interdisciplinary field. The inclusion of eight major databases spanning health sciences, psychology, and technology domains reflects the multidisciplinary nature of AI-enhanced interventions and minimizes the risk of missing relevant studies published outside traditional gerontology journals.

The use of Covidence for study management represents a methodological strength, providing transparent documentation of the screening process and facilitating collaboration between reviewers while maintaining independence in decision-making. The platform’s automated conflict detection and resolution tracking enhances the reliability and reproducibility of the selection process. The comprehensive risk of bias assessment using validated tools appropriate to each study design ensures that evidence quality is systematically evaluated, informing both the interpretation of findings and recommendations for practice and research.

Several challenges are anticipated in conducting this review. The heterogeneity expected across studies in terms of AI technologies employed, reminiscence therapy protocols, outcome measures, and population characteristics may limit opportunities for quantitative synthesis. The nascent state of the field suggests that many studies may be exploratory or pilot investigations with small sample sizes, potentially limiting the strength of conclusions. The rapid pace of technological advancement means that findings may not fully reflect the current capabilities of AI systems, highlighting the importance of considering temporal trends in the analysis.

The protocol’s broad inclusion criteria, encompassing various study designs and populations across the cognitive spectrum, reflects a deliberate decision to capture the full scope of current evidence while acknowledging that this may increase heterogeneity. This inclusive approach is justified given the exploratory nature of the field and the value of understanding implementation experiences and user perspectives alongside effectiveness data. The inclusion of qualitative and mixed-methods studies will provide crucial insights into acceptability, feasibility, and implementation factors that purely quantitative approaches might miss.

Ethical considerations permeate this research area, particularly regarding vulnerable populations with cognitive impairment. The review will critically examine how included studies address informed consent, data privacy, and the preservation of human dignity in technology-mediated interventions. The synthesis will highlight best practices and identify gaps in ethical frameworks, contributing to the development of guidelines for responsible implementation of AI in therapeutic contexts for older adults.

The review’s findings will have important implications for multiple stakeholders. For clinicians and care providers, the synthesis will offer evidence-based guidance on selecting and implementing AI-enhanced reminiscence interventions, including insights into which technologies show promise for specific populations and outcomes. For technology developers, the review will identify design requirements, user needs, and implementation challenges that should inform future innovation. For policymakers, evidence regarding effectiveness, cost-effectiveness, and implementation requirements will support informed decision-making about resource allocation and regulatory frameworks.

The protocol acknowledges several limitations that may influence the review’s scope and conclusions. The focus on published literature may miss innovations in commercial applications that have not undergone empirical evaluation. The interdisciplinary nature of the field presents challenges for comprehensive searching, despite extensive database coverage and supplementary search strategies. The quality of the review’s findings will ultimately depend on the quality of available primary studies, which may be limited given the field’s early stage of development.

This protocol contributes to the broader discourse on AI in healthcare by examining a specific application that balances technological innovation with human-centered care principles. The systematic approach to synthesizing evidence will provide a foundation for understanding how AI can augment rather than replace therapeutic relationships, offering lessons applicable to other domains of AI application in health and social care. The review will also contribute to theoretical understanding of reminiscence processes by examining how AI-mediated approaches align with or challenge existing frameworks of life review and autobiographical memory in aging.

## Data Availability

All data produced in the present work are contained in the manuscript

## References

1. World Health O. Global report on ageism. 2021.

2. Woods B, O’Philbin L, Farrell EM, Spector AE, Orrell M. Reminiscence therapy for dementia. Cochrane Database Syst Rev. 2018;3(3):Cd001120.

3. Butler RN. The life review: an interpretation of reminiscence in the aged. Psychiatry. 1963;26:65–76.

4. Westerhof GJ, Bohlmeijer ET. Celebrating fifty years of research and applications in reminiscence and life review: state of the art and new directions. J Aging Stud. 2014;29:107–14.

5. Pinquart M, Forstmeier S. Effects of reminiscence interventions on psychosocial outcomes: a meta-analysis. Aging Ment Health. 2012;16(5):541–58.

6. Cotelli M, Manenti R, Zanetti O. Reminiscence therapy in dementia: A review. Maturitas. 2012;72(3):203–5.

7. Lazar A, Thompson H, Demiris G. A systematic review of the use of technology for reminiscence therapy. Health education & behavior. 2014;41(1_suppl):51S–61S.

8. Chen K, Chan AH. Gerontechnology acceptance by elderly Hong Kong Chinese: a senior technology acceptance model (STAM). Ergonomics. 2014;57(5):635–52.

9. Zhu A, Luximon Y, editors. Rethinking Dementia in Aging with Generative AI Technologies: A Preliminary Conceptual Framework 2025: SAGE Publications Sage CA: Los Angeles, CA.

10. Jin Y, Cai W, Chen L, Zhang Y, Doherty G, Jiang T, editors. Exploring the design of generative AI in supporting music-based reminiscence for older adults 2024.

11. García-Betances RI, Arredondo Waldmeyer MT, Fico G, Cabrera-Umpiérrez MF. A succinct overview of virtual reality technology use in Alzheimer’s disease. Front Aging Neurosci. 2015;7:80.

12. Shankar R, Bundele A, Mukhopadhyay A. A Systematic Review of Natural Language Processing Techniques for Early Detection of Cognitive Impairment. Mayo Clin Proc Digit Health. 2025;3(2):100205.

13. Schardt C, Adams MB, Owens T, Keitz S, Fontelo P. Utilization of the PICO framework to improve searching PubMed for clinical questions. BMC Med Inform Decis Mak. 2007;7:16.

14. Sterne JAC, Savović J, Page MJ, Elbers RG, Blencowe NS, Boutron I, et al. RoB 2: a revised tool for assessing risk of bias in randomised trials. Bmj. 2019;366:l4898.

15. Sterne JA, Hernán MA, Reeves BC, Savović J, Berkman ND, Viswanathan M, et al. ROBINS-I: a tool for assessing risk of bias in non-randomised studies of interventions. Bmj. 2016;355:i4919.

16. Long HA, French DP, Brooks JM. Optimising the value of the critical appraisal skills programme (CASP) tool for quality appraisal in qualitative evidence synthesis. Research Methods in Medicine & Health Sciences. 2020;1(1):31–42.

17. Guyatt GH, Oxman AD, Vist GE, Kunz R, Falck-Ytter Y, Alonso-Coello P, et al. GRADE: an emerging consensus on rating quality of evidence and strength of recommendations. Bmj. 2008;336(7650):924–6.

18. Higgins JP, Thompson SG, Deeks JJ, Altman DG. Measuring inconsistency in meta-analyses. Bmj. 2003;327(7414):557–60.

19. Thomas J, Harden A. Methods for the thematic synthesis of qualitative research in systematic reviews. BMC medical research methodology. 2008;8(1):45.

